# Determinants of total and inhaled allergen-specific immunoglobulin E in the middle-aged and elderly population

**DOI:** 10.64898/2026.05.12.26352742

**Authors:** M. Al Fatly, S. Leonard, P.L.A van Daele, G. Helleman, E. Torabi-Azhandeh, L. Lahousse, S. Veenbergen, L. Chaker

## Abstract

**Backgroun:** The determinants of immunoglobulin E (IgE) remain poorly understood in older adults, a population with an increasing burden of chronic diseases. Identifying IgE’s determinants may improve its clinical interpretation in the evaluation of allergic and IgE-related conditions.

**Objective:** To investigate age, sex, smoking, alcohol, body mass index (BMI), corticosteroid use, and season as potential determinants of total IgE (tIgE) and inhaled allergen-specific IgE (sIgE).

**Methods:** Using Rotterdam Study data, we investigated the determinants of tIgE and sIgE using multivariable linear regression. Longitudinal changes and the effects of corticosteroids were assessed with linear mixed models.

**Results:** We included 8769 participants, of which 478 had repeated IgE measurements. Age showed a U-shaped relationship with tIgE and L-shaped relationship with sIgE (both p<0.001). Women had lower tIgE (OR [95%CI]: 0.69 [0.65-0.74]), whereas current smokers (1.34 [1.23-1.46]), higher BMI (1.01 [1.01-1.02]), topical corticosteroid users (1.27 [1.07-1.50]) and inhaled corticosteroid users (1.93 [1.64-2.26]) showed higher tIgE. Women (0.96 [0.92-1.00]), former smokers (0.87 [0.83-0.91]) and current smokers (0.72 [0.68-0.76]) had lower sIgE, whereas topical corticosteroid users (1.20 [1.07-1.35]) and inhaled corticosteroid users (1.20 [1.07-1.35]) showed higher sIgE. Over time, tIgE and sIgE decreased (p<0.001) but did not significantly change after corticosteroid use.

**Conclusion:** We identified age, sex, smoking, BMI, season and topical and inhaled corticosteroids as determinants of tIgE and sIgE. Incorporating these determinants may improve IgE’s clinical interpretation for the diagnosis and management of allergic and IgE-related conditions. Future research should investigate how these determinants shape IgE’s relationship with chronic diseases in aging populations.

## Introduction

Immunoglobulin E (IgE) plays a key role in hypersensitivity reactions and immune responses against parasites and venom (1). While total IgE (tIgE) levels can show general IgE abnormalities, allergen-specific IgE (sIgE) levels can confirm allergen-specific sensitization (2, 3). Both can aid in diagnosing and managing allergic and other IgE-related conditions (2-4). Beyond IgE’s conventionally established roles, there is increasing evidence linking tIgE and sIgE to chronic diseases, including cancer (5-17), cardiovascular disease (18) and neurodegenerative disorders (19), potentially through chronic inflammatory mechanism (5-19).

Although tIgE and sIgE are key clinical biomarkers with a potential role in chronic disease, their determinants remain poorly understood. This knowledge gap is especially evident in the general middle-aged and elderly population, where the burden of chronic diseases is increasing. Non-modifiable factors like age, sex and season, and modifiable factors like smoking, alcohol consumption, body mass index (BMI) and corticosteroid use, have previously been linked to altered tIgE and sIgE levels (20-25). However, most studies were limited by small sample sizes and a lack of generalizability from exclusively including children or atopic individuals (26, 27). Additionally, inconsistencies across studies in the measurement of tIgE and sIgE hinder comparisons.

Given previous limitations, there remains a gap in knowledge on the determinants of tIgE and sIgE in middle-aged and older adults. Identifying these determinants may improve the clinical interpretation of tIgE and sIgE in the diagnosis and management of allergic and IgE-related conditions. Additionally, it is essential for future research on the potential causal relationships of tIgE and sIgE with chronic diseases. Therefore, we investigated age, sex, smoking, alcohol use, BMI, season and corticosteroid use as potential determinants of tIgE and sIgE in a cohort of general middle-aged and older adults. We also examined the change in tIgE and sIgE levels over time, and the dose-response effects of corticosteroid use.

## 2. Methods

### 2.1 Study design

We used data from the Rotterdam Study (RS), a large, ongoing, population-based cohort study initiated in 1990, including middle-aged and elderly residents of Ommoord, Rotterdam, The Netherlands. The focus of the RS is primarily on cardiovascular, endocrine, hepatic, neurological, ophthalmic, psychiatric, dermatological, otolaryngological, locomotor, and respiratory diseases. The RS has been approved by the Medical Ethics Committee of the Erasmus MC (registration number MEC 02.1015) and the Dutch Ministry of Health, Welfare and Sport (Population Screening Act WBO, license number 1071272-159521-PG). The Personal Registration Data collection of the RS is filed with the Erasmus MC Data Protection Officer under registration number EMC1712001. The RS is listed in the Netherlands National Trial Register and in the WHO International Clinical Trials Registry Platform under shared catalogue number NTR6831. All participants provided written informed consent to participate in the study and to have their information obtained from treating physicians. The aims and study design of the RS have been previously published in detail (28).

### 2.2 Study population

We included cross-sectional data of participants from three independent RS cohorts (RSI-1, RSII-1 and RSIII-1). Data from home interviews, research center examinations and blood samples were collected during participants’ first visit between 1989-1993 (RSI-1), 2000-2001 (RSII-1), and 2006-2008 (RSIII-1). Additionally, we selected a random sample from RSIII-1 where second measurements of serum tIgE and sIgE were conducted during the participants’ second visit, between 2012-2014 (RSIII-2).

### 2.3 Determinants

Based on previous literature, biological plausibility and data availability, we extracted age, sex, smoking status, alcohol consumption, BMI, season and corticosteroid use, including topical (ATC code D07), inhaled (ATC codes RB03ba, RB0ak, RB0al) and oral (ATC code H02) as potential determinants. Anthropometric data was collected during physical examination. BMI was calculated as weight/height^2^ (kg/m^2^). Season was based on blood sampling dates and categorized as spring, summer, autumn, winter.

We extracted smoking status, alcohol consumption and topical corticosteroid use from home interviews. Smoking status was defined as current, former and never. Information on alcohol consumption was available in grams per day and categorized by WHO guidelines as 0-10 g/day (mild), 10-20 g/day (moderate), >20 g/day (heavy). Topical corticosteroid use was dichotomized (yes/no). We extracted information on inhaled and oral corticosteroids from medication dispensing data gathered through automated linkage of pharmacy records, where current use at the blood sampling date was reported. For the longitudinal analysis, we extracted the defined daily dose (DDD), a standardized average daily dose for a medication’s main indication (29). We calculated each individual’s cumulative DDD per year (cDDD/year) from the DDDs between their first and second measurement. Corticosteroid use was categorized as none (0), moderate (<180), or frequent (>180).

### 2.4 Assessment of tIgE and sIgE

Serum tIgE and sIgE levels were measured between December 2023 and November 2024 in Erasmus MC using NOVEOS (Hycor, USA), a fully automated, high-throughput chemiluminescent immunoassay system (30). The detection range was 5-5000 kU/L (kilounits per liter) for tIgE. Total IgE >100 kU/L was considered elevated. The molecular SX01 IgE test was used to determine IgE sensitization to inhaled allergens, consisting of a balanced mix of recombinant allergen molecules (Der p 1/2, Can f 1/2/3/5, Fel d 1, Bet v 1, Phl p 1, Art v 1). Standard Arbitrary Units (SAU) ≥1 was considered positive for sIgE sensitization. The NOVEOS immuno-analyzer minimizes biotin and cellulose-based CCD interference by using paramagnetic microparticles and a non-competitive indirect immunoassay design, improving assay performance (31). In-house validation revealed intra- and inter-assay variation coefficients of 4.0% and 5.2%.

### 2.5 Statistical analysis

We assessed the cross-sectional relationships between the determinants and tIgE and sIgE using multivariable linear regression models. We included age, sex, smoking status, alcohol consumption, BMI, season, and topical, inhaled, and oral corticosteroids as covariates. For the main analysis, we excluded tIgE values below the detection limit and zero values of sIgE, and log-transformed tIgE and sIgE to get normally distributed residuals. We assessed nonlinearity of age through restricted cubic splines with four knots at the 5^th^, 35^th^, 65^th^ and 95^th^ percentiles. As sensitivity analyses, we investigated the age-sex interaction and assessed batch and cohort variance through linear regression, stratification and linear mixed models. Additionally, we performed Tobit regression as a sensitivity analysis to investigate tIgE’s determinants when including and accounting for values below the detection limit within the analysis. By comparing Tobit regression results to linear regression results, we investigated potential issues of selection bias that could arise from excluding values below the detection limit in analysis.

For the longitudinal analyses, we assessed changes in tIgE and sIgE between the first and second measurements. For this, we used linear mixed models adjusted for sex, smoking, alcohol consumption, BMI and corticosteroid use. We investigated dose-response effects of inhaled and oral corticosteroids on tIgE and sIgE in separate linear mixed models, adjusted for age, sex, smoking, alcohol consumption and BMI. For both analyses, we log-transformed tIgE and sIgE to obtain normally distributed residuals. To preserve statistical power, we did not exclude tIgE values below the detection limit. We performed an inverse probability weighting (IPW) sensitivity analysis to assess potential selection bias in the longitudinal sample.

We performed multiple imputations by chained equations using 10 imputations and 20 iterations to impute missing values. The percentage of missing values was <2% for all determinants except alcohol consumption (8.4%). We z-standardized tIgE and sIgE for comparison in all analyses. All statistical analyses were conducted in R statistical software versions 4.4.1 using mice (32), nnet (33), censReg (34) and lme4 (35).

## 3 Results

### 3.1 Study population characteristics

We included 8679 participants (mean±SD age 64.2±9.8 years; 56.9% female) in the cross-sectional analysis (Table 1). Of 8671 participants with tIgE measurements, 7751 had detectable values and were included in the tIgE analysis. For sIgE, 8662 participants had measurements and were included in the sIgE analysis. The median tIgE was 24.3 kU/L and the median sIgE was 0.12 SAU. Furthermore, 19.3% had tIgE levels >100 kU/L and 16.5% had sIgE levels >1 SAU. The determinants of tIgE and sIgE are depicted in Figures 1 and 2, respectively, and the effect estimates per determinant in Table 2.

**Table 1:**
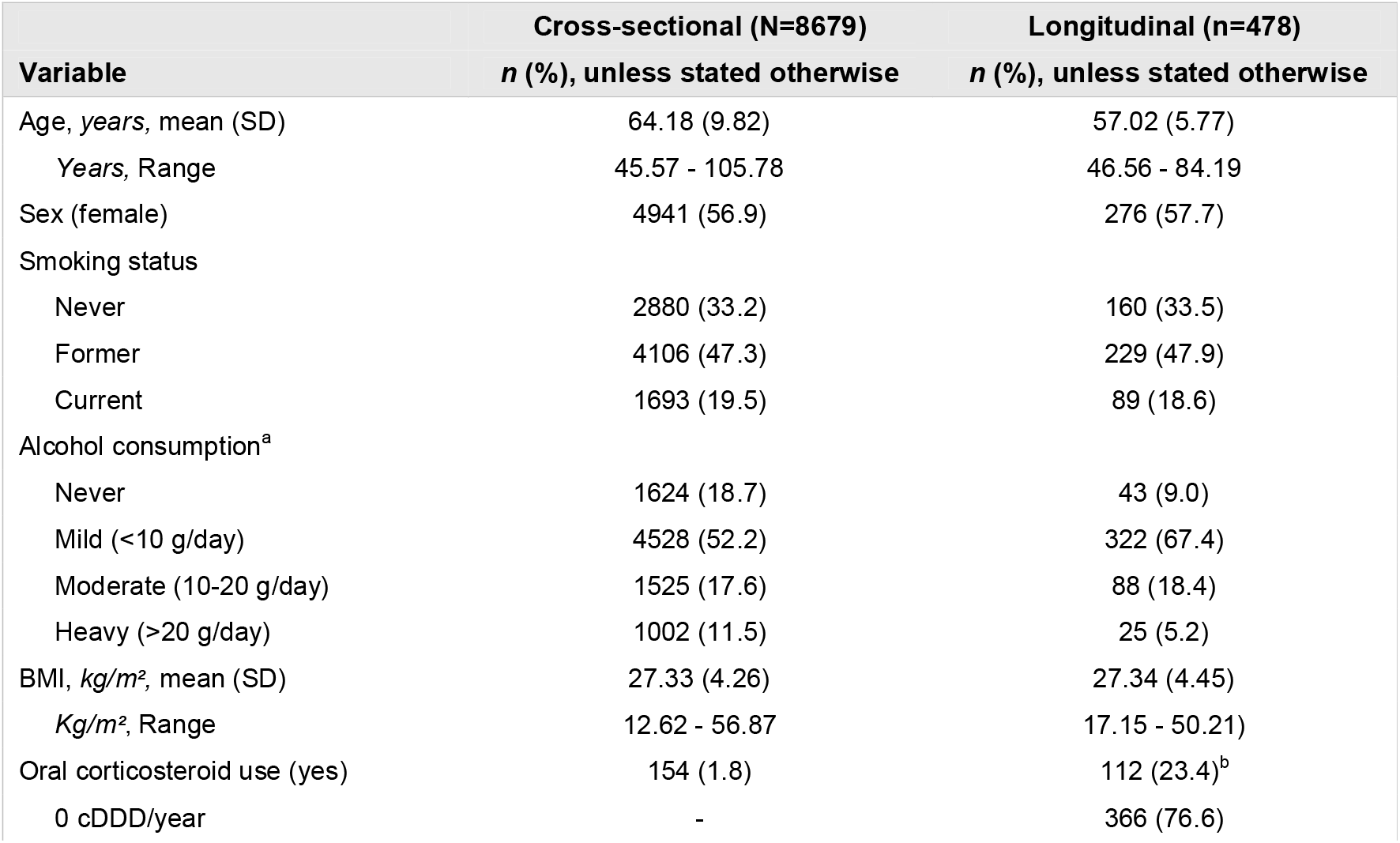
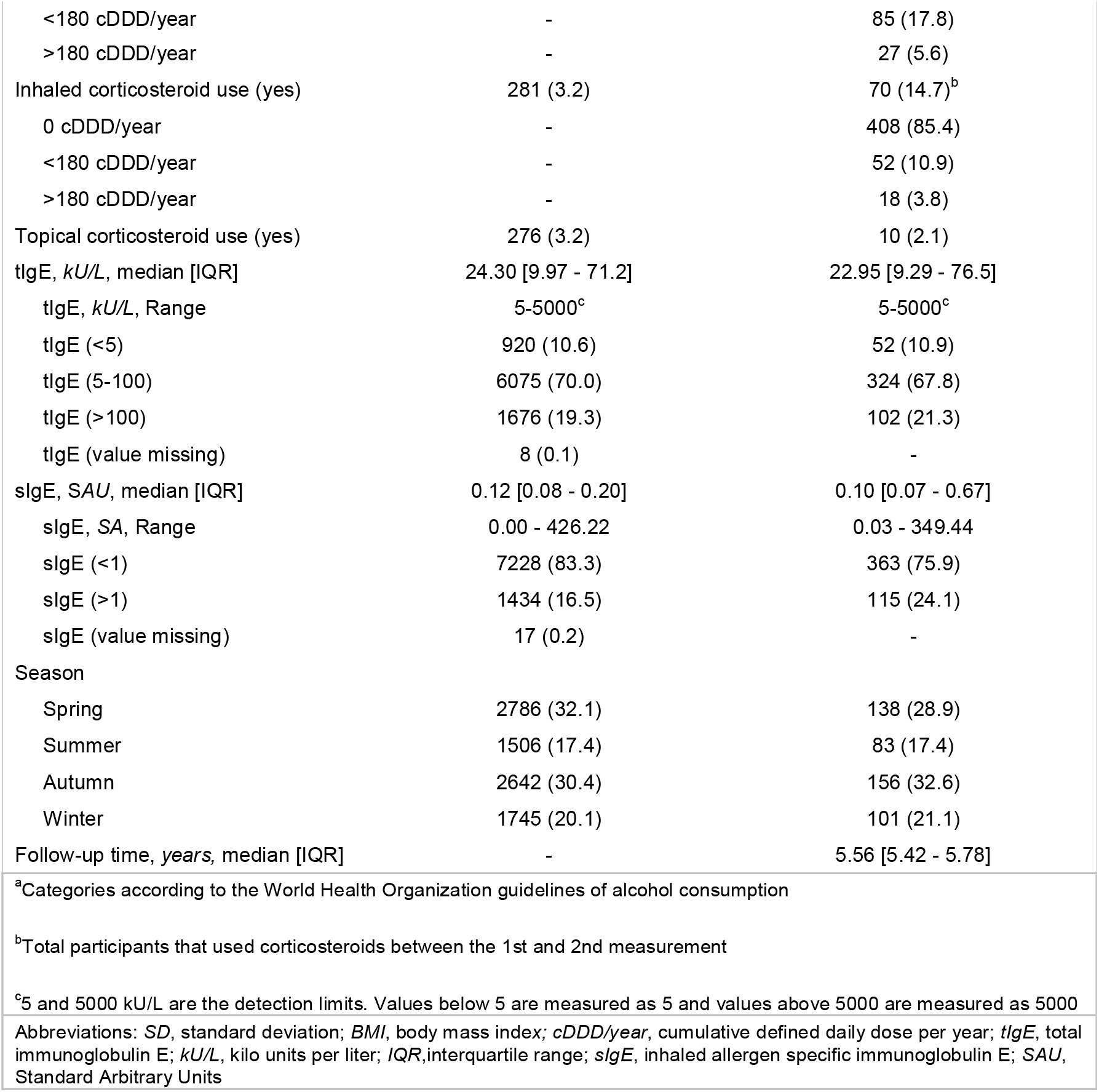
Population characteristics of the cross-sectional and longitudinal sample.

| Variable | Cross-sectional (N=8679)<br>*n* (%), unless stated otherwise | Longitudinal (n=478)<br>*n* (%), unless stated otherwise |
| --- | --- | --- |
| Age, years, mean (SD) | 64.18 (9.82) | 57.02 (5.77) |
| Years, Range | 45.57 - 105.78 | 46.56 - 84.19 |
| Sex (female) | 4941 (56.9) | 276 (57.7) |
| Smoking status |  |  |
| Never | 2880 (33.2) | 160 (33.5) |
| Former | 4106 (47.3) | 229 (47.9) |
| Current | 1693 (19.5) | 89 (18.6) |
| Alcohol consumption<sup>a</sup> |  |  |
| Never | 1624 (18.7) | 43 (9.0) |
| Mild (<10 g/day) | 4528 (52.2) | 322 (67.4) |
| Moderate (10-20 g/day) | 1525 (17.6) | 88 (18.4) |
| Heavy (>20 g/day) | 1002 (11.5) | 25 (5.2) |
| BMI, kg/m<sup>2</sup>, mean (SD) | 27.33 (4.26) | 27.34 (4.45) |
| kg/m<sup>2</sup>, Range | 12.62 - 56.87 | 17.15 - 50.21) |
| Oral corticosteroid use (yes) | 154 (1.8) | 112 (23.4)<sup>b</sup> |
| 0 cDDD/year | - | 366 (76.6) |
| <180 cDDD/year | - | 85 (17.8) |
| >180 cDDD/year | - | 27 (5.6) |
| Inhaled corticosteroid use (yes) | 281 (3.2) | 70 (14.7)<sup>b</sup> |
| 0 cDDD/year | - | 408 (85.4) |
| <180 cDDD/year | - | 52 (10.9) |
| >180 cDDD/year | - | 18 (3.8) |
| Topical corticosteroid use (yes) | 276 (3.2) | 10 (2.1) |
| tlgE, kU/L, median [IQR] | 24.30 [9.97 - 71.2] | 22.95 [9.29 - 76.5] |
| tlgE, kU/L, Range | 5-5000<sup>c</sup> | 5-5000<sup>c</sup> |
| tlgE (<5) | 920 (10.6) | 52 (10.9) |
| tlgE (5-100) | 6075 (70.0) | 324 (67.8) |
| tlgE (>100) | 1676 (19.3) | 102 (21.3) |
| tlgE (value missing) | 8 (0.1) | - |
| slgE, SAU, median [IQR] | 0.12 [0.08 - 0.20] | 0.10 [0.07 - 0.67] |
| slgE, SA, Range | 0.00 - 426.22 | 0.03 - 349.44 |
| slgE (<1) | 7228 (83.3) | 363 (75.9) |
| slgE (>1) | 1434 (16.5) | 115 (24.1) |
| slgE (value missing) | 17 (0.2) | - |
| Season |  |  |
| Spring | 2786 (32.1) | 138 (28.9) |
| Summer | 1506 (17.4) | 83 (17.4) |
| Autumn | 2642 (30.4) | 156 (32.6) |
| Winter | 1745 (20.1) | 101 (21.1) |
| Follow-up time, years, median [IQR] | - | 5.56 [5.42 - 5.78] |
| <sup>a</sup>Categories according to the World Health Organization guidelines of alcohol consumption |  |  |
| <sup>b</sup>Total participants that used corticosteroids between the 1st and 2nd measurement |  |  |
| <sup>c</sup>5 and 5000 kU/L are the detection limits. Values below 5 are measured as 5 and values above 5000 are measured as 5000 |  |  |
| Abbreviations: *SD*, standard deviation; *BMI*, body mass index; *cDDD/year*, cumulative defined daily dose per year; *tlgE*, total immunoglobulin E; *kU/L*, kilo units per liter; *IQR*, interquartile range; *slgE*, inhaled allergen specific immunoglobulin E; *SAU*, Standard Arbitrary Units |  |  |

**Figure 1:**
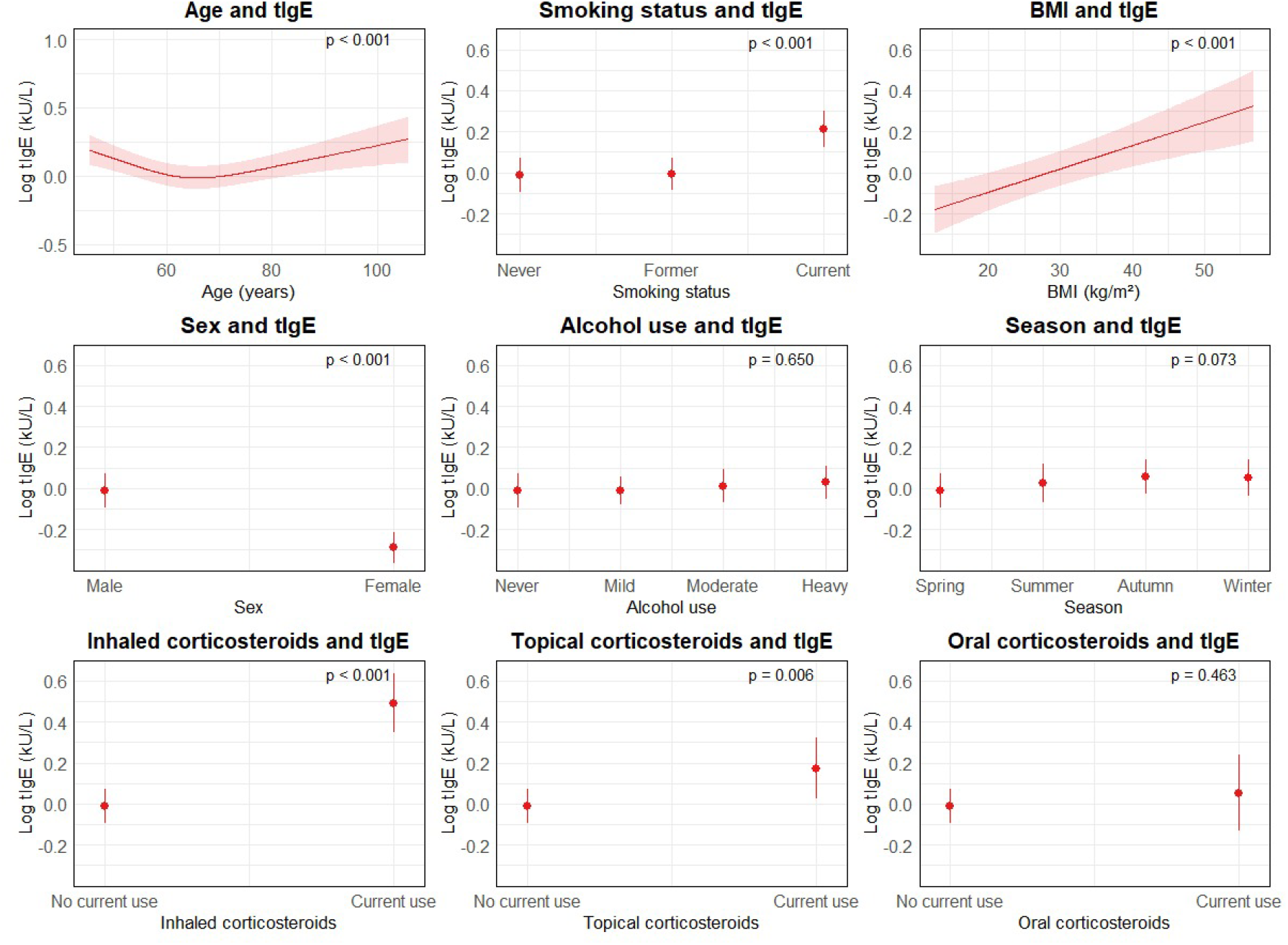
Effect plots of the determinants of tIgE. **Abbreviations**: *tIgE*, total immunoglobulin E; *kU/L*, kilo units per liter; *BMI*, body mass index **Description**: Figure 1 shows the effect plots of the relationships between the potential determinants and tIgE, adjusted for age, sex, smoking, alcohol consumption, BMI, season and topical, inhaled and oral corticosteroids. From left to right, top to bottom, the plots are presented for age, sex, smoking, alcohol consumption, BMI, and inhaled, topical and oral corticosteroids. The tIgE values were log-transformed after exclusion of the values below the detection limit. The effect estimates are standardized through z-score standardization and plotted as beta coefficient per standard deviation.

**Figure 2:**
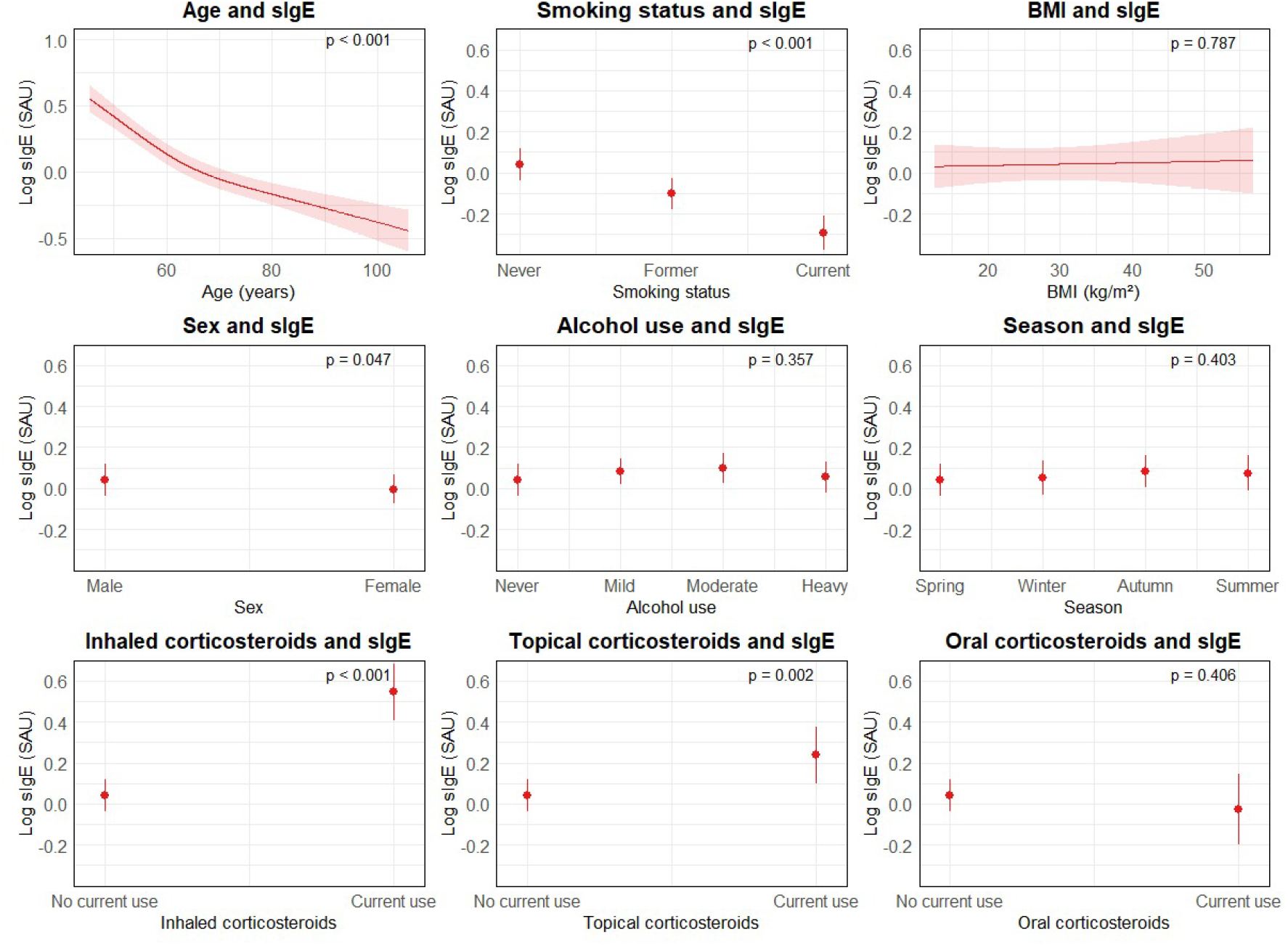
Effect plots of the determinants of sIgE. **Abbreviations**: *sIgE*, inhaled allergen specific immunoglobulin E; *SAU*, Standard Allergen Unit; *BMI*, body mass index; *BMI*, body mass index **Description**: Figure 2 shows the effect plots of the relationships between the potential determinants and sIgE, adjusted for age, sex, smoking, alcohol consumption, BMI, season and topical, inhaled and oral corticosteroids. From left to right, top to bottom, the plots are presented for age, sex, smoking, alcohol consumption, BMI, and inhaled, topical and oral corticosteroids. The sIgE values were log-transformed after exclusion of zero values. The effect estimates are standardized through z-score standardization and plotted as beta coefficient per standard deviation.

### 3.2 Determinants of tIgE

Figure 1 depicts the relationships of each determinant with tIgE levels. We found a nonlinear, U-shaped relationship between age and tIgE (p for nonlinearity <0.001). Until approximately 70 years of age, older age was associated with lower tIgE, while after 70 years of age, older age was associated with higher tIgE values. Women had lower tIgE values than men (OR [95%CI]: 0.69 [0.65-0.74]). Although the age-sex interaction was not statistically significant, the plotted relationship suggests that in men, higher age is associated with higher tIgE values than in women (p=0.080, Supplementary Figure S1). Current smokers had higher tIgE values compared to never smokers (OR [95%CI]: 1.34 [1.23-1.46]), but there was no difference between former and never smokers (OR [95%CI]: 1.01 [0.94-1.08]). Alcohol consumption was not associated with tIgE levels (p=0.183). Higher BMI was associated with higher tIgE values (OR [95%]: 1.01 [1.01-1.02]). Topical and inhaled corticosteroid use were both associated with higher tIgE. This difference was largest for inhaled corticosteroid use (OR [95%CI] 1.93 [1.64-2.26]), followed by topical corticosteroid use 1.27 [1.07-1.50]. In contrast, tIgE values did not differ significantly with oral corticosteroid use (OR [95%CI]: 1.09 [0.87-1.36]). For season, tIgE values were higher in individuals measured in autumn and winter compared to spring (OR [95%CI], respectively for autumn and winter: 1.09 [1.02-1.17] and 1.09 [1.00-1.18]). There was no difference between summer and spring (OR [95%CI]: 1.05 [0.96-1.14]).

### 3.3 Determinants of sIgE

Figure 2 shows the relationships between the determinants and sIgE levels. There was a nonlinear, L-shaped relationship between age and sIgE (p for nonlinearity <0.001). Women had significantly lower sIgE levels than men (OR [95%CI]: 0.96 [0.92-1.00]). The age-sex interaction was nonsignificant (p for interaction: 0.196, Supplementary Figure S1). Former and current smokers had significantly lower sIgE levels than never smokers, with current smokers showing the lowest levels (OR [95%CI] for former and current smokers, respectively: 0.87 [0.83-0.91] and 0.72 [0.68-0.76]). Furthermore, both topical and inhaled and corticosteroid use were significantly associated with higher sIgE levels (OR [95%CI] for topical and inhaled corticosteroid use, respectively: 1.20 [1.07-1.35] and 1.64 [1.47-1.84]), while oral corticosteroid use was not associated with sIgE levels (OR [95%CI]: 0.94 [0.80-1.10]). Alcohol consumption (p=0.264), BMI (p=0.703) and season (p=0.422) were not significantly associated with sIgE levels.

### 3.4 Sensitivity analyses

There was no significant relationship between tIgE levels and cohorts or batches. For sIgE, cohort stratification did not meaningfully change results. Although sIgE values were significantly higher in batches 10 and 11 compared to reference batch 1, excluding those batches did not meaningfully change results and the linear mixed model showed no significant batch differences (Supplementary Figure S2). For tIgE, the Tobit and linear regression effect estimates were similar, indicating no major issue of selection bias from excluding tIgE values below the detection limit in the linear regression (Table 2).

### 3.5 Change in tIgE and sIgE over time

In the longitudinal analysis, we included 478 participants with two measurements of both tIgE and sIgE. The median [IQR] time between the measurements was 5.56 [5.42-5.78] years (Table 1). Mean age was lower than in the total study population (mean±SD: 57.02±5.77 vs 64.18±9.82), but similar to RSIII-1 (mean±SD: 57.82±8.42). Other characteristics were also similar to RSIII-1 (Supplementary Table S1). Both tIgE and sIgE decreased over time (respectively, OR [95%CI]: 0.94 [0.91-0.98] and 0.94 [0.92-0.96], Figure 3). Despite the difference in age distribution between the longitudinal sample and total study population, applying IPW did not change the results meaningfully, indicating no major issue of selection bias.

**Figure 3:**
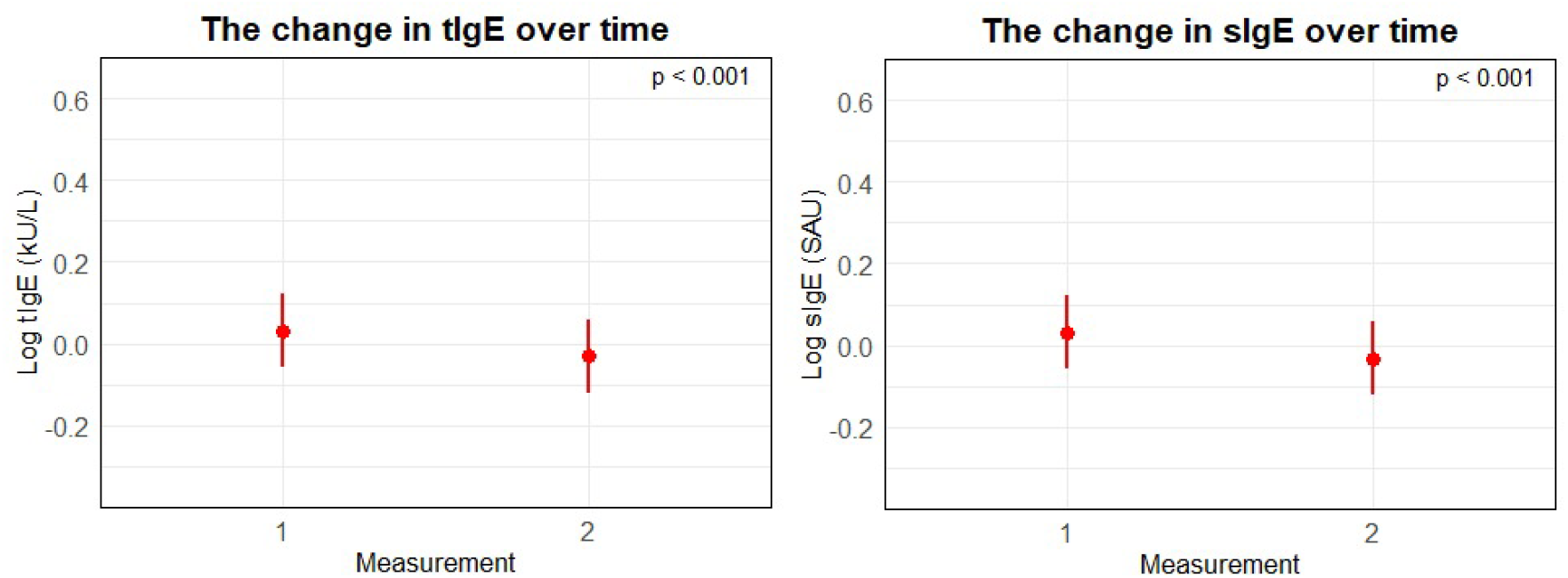
The change in tIgE and sIgE over time. **Abbreviations**: *tIgE*, total immunoglobulin E; *kU/L*, kilo units per liter; *sIgE*, inhaled allergen specific immunoglobulin E; *SAU*, Standard Arbitrary Units; **Description**: Figure 3 shows the effect plots of the change in tIgE (left) and sIgE (right) over time, adjusted for age, sex, smoking, alcohol consumption, BMI, season and topical, oral and inhaled corticosteroids. Both tIgE and sIgE are log-transformed. The effect estimates are standardized through z-score standardization and plotted as beta coefficient per standard deviation.

### 3.6 Change in tIgE and sIgE with corticosteroids

Between the first and second measurements, 112 participants used oral corticosteroids, with 85 participants using <180 cDDD/year and 27 using >180 cDDD/year. Seventy participants used inhaled corticosteroids, with 52 using <180 cDDD/year and 18 using >180 cDDD/year. Figure 4 depicts the change in tIgE and sIgE after inhaled and oral corticosteroid use and Table 2 presents the effect estimates. There was no significant change in tIgE or sIgE after inhaled or oral corticosteroid use, although figure patterns suggest that tIgE and sIgE levels may increase with higher cDDD/year of inhaled corticosteroids and decrease with higher cDDD/year of oral corticosteroids.

**Figure 4:**
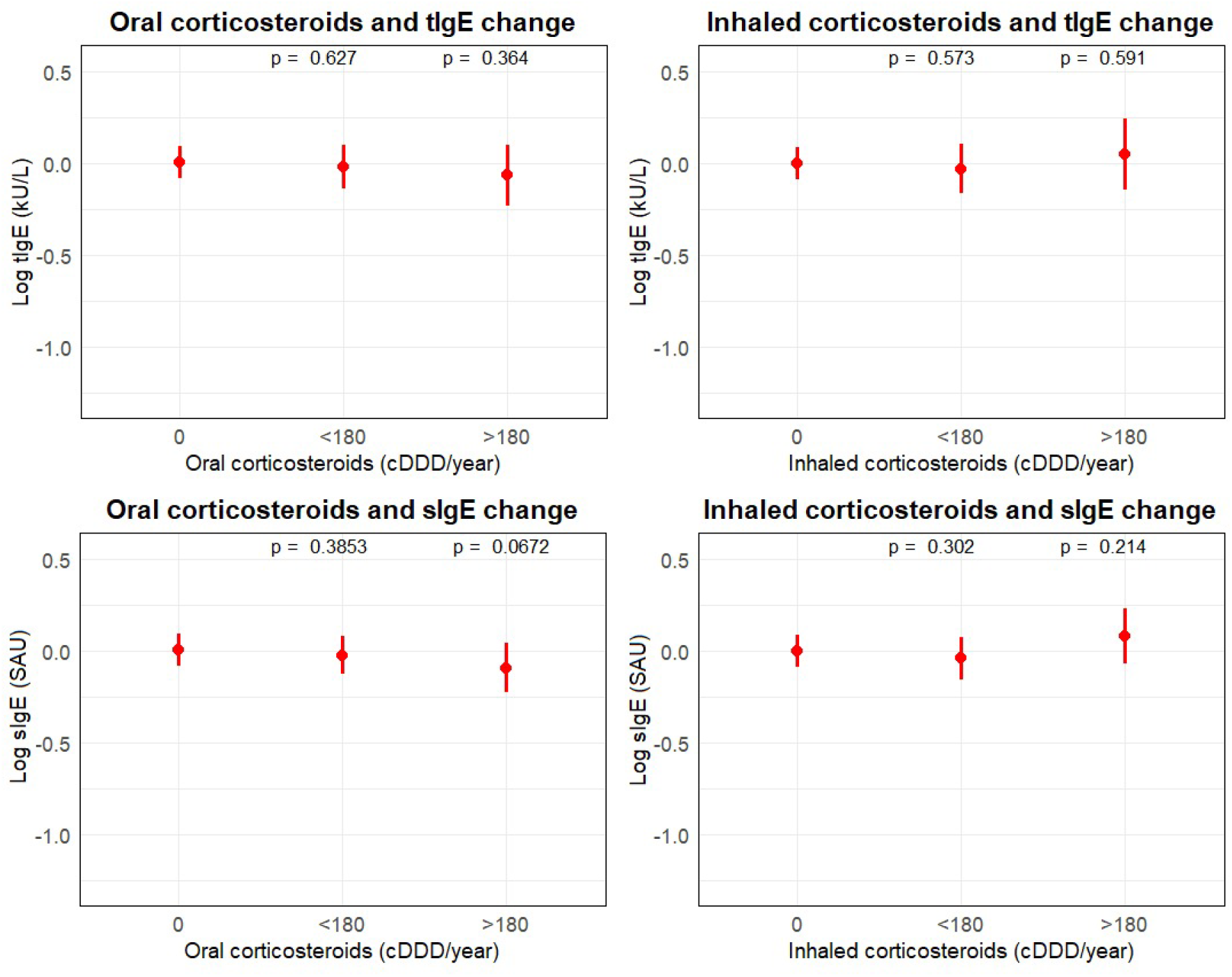
The change in tIgE and sIgE after oral and inhaled corticosteroid use. **Abbreviations**: *tIgE*, total immunoglobulin E; *kU/L*, kilo units per liter; *cDDD/year*, cumulative defined daily dose per year; *sIgE*, inhaled allergen specific immunoglobulin E; *SAU*, Standard Arbitrary Units **Description**: Figure 4 shows the effect plots of the change in tIgE and sIgE after corticosteroid use, per category of cDDD/year. The top left and right plots show the change in tIgE after, respectively, oral and inhaled corticosteroids. The bottom left and right plots show the change in sIgE after, respectively, oral and inhaled corticosteroids. The corticosteroid types were included one by one per model and each model was adjusted for age, sex, smoking, alcohol consumption, BMI and topical corticosteroid use. Both tIgE and sIgE are log-transformed. The effect estimates are standardized through z-score standardization and plotted as beta coefficient per standard deviation.

## 4 Discussion

In this large population-based cohort of general middle-aged and older adults, we investigated the determinants of tIgE and sIgE. We found a U-shaped relationship between tIgE and age. Female sex was associated with lower tIgE, and smoking, higher BMI, autumn and winter, and topical and inhaled corticosteroid use were associated with higher tIgE. For sIgE, there was an L-shaped relationship with age. Smoking and female sex were associated with lower sIgE, while topical and inhaled corticosteroid use were associated with higher sIgE. Longitudinally, both tIgE and sIgE decreased over time, but we found no dose-response effect of inhaled or oral corticosteroids.

First, age demonstrated a nonlinear association with both tIgE and sIgE. Higher age was associated with lower tIgE levels in ages below 70, and higher tIgE levels after 70 years. These results align with previous findings showing a U-shaped relationship between age and tIgE (36, 37). Although some previous studies showed inverse association across all ages (37-39), or no association (25, 40-42), these studies mainly included smaller samples, younger individuals, analyzed categories of age instead of continuous age, and did not assess nonlinearity. As for sIgE, an increase in age was associated with lower sIgE levels. This association was more outspoken in younger ages. Methodological differences limit direct comparisons, as one study investigated dichotomous sIgE sensitization (36), and others investigated sIgE by allergen in allergic individuals (43, 44). Nevertheless, these studies found less sensitization in older age, supporting our findings of lower sIgE in older age. Furthermore, we found that tIgE and sIgE decreased over time. To our knowledge, this is the first study (44), to show the change in tIgE and sIgE over time in a general middle-aged and elderly population. Mechanistically, the tIgE patterns may be explained by age-related T- and B-cell remodeling, including reduced IL-4 production and impaired isotype switching to IgE (45). For sIgE, long-term exposure to antigens throughout life could be responsible for immune tolerance against those antigens and therefore lower sIgE levels in older ages (45). A previous systematic review showed lower IgM and higher IgA levels in older individuals (46) and another study reported a nonlinear pattern for IgG similar to IgE (47). Hence, the associations of tIgE and sIgE with age may reflect broader age-related immune remodeling, as described in previous literature (36, 43).

In line with previous studies, we found lower tIgE and sIgE levels in women compared to men (25, 38, 39, 48, 49). While we investigated sIgE for a mixture of inhaled allergens, previous studies focused on sIgE per specific allergen and showed a sex difference for some allergens, such as house dust mite, but not all (36, 43, 50). Although not statistically significant, the age-sex interaction plot suggested that in men older than 70, older age may be associated with a more outspoken increase in tIgE levels than in women older than 70. We found no significant age-sex interaction for sIgE. To our knowledge, age-sex interaction has not been previously investigated for IgE. A possible explanation for the sex differences is the influence of sex hormones, which may contribute to variations in immune responses, antibody production and isotype switching (48, 51). Although not well investigated yet, sex hormones may play a role in translating IgE sensitization into clinical manifestation of atopic and allergic symptoms. This could explain the paradoxical higher prevalence of allergy and atopy in women compared to men, despite their lower tIgE levels (52).

Current smokers had higher tIgE compared to never smokers. This aligns with previous studies conducted in smaller and specific populations (53-56) and may result from IL-4-mediated dysregulated isotype switching triggered by inflammation (55, 57, 58). Conversely, former and current smokers had lower sIgE. This could reflect reverse causation, where individuals with allergic tendencies are less likely to smoke (50, 59). Interestingly, alcohol was not significantly associated with tIgE or sIgE. Some previous studies found heavy alcohol consumption to be associated with higher tIgE (25, 60, 61), and higher sIgE for some specific allergens like mites, but not all (61, 62). Possible reasons for these inconsistent findings could be investigating non-atopic individuals only, categorizing alcohol consumption differently, adjusting for different covariates and comparing geometric means. Additionally, the studies on sIgE mainly focused on sIgE per specific allergens, while we investigated a mixture of inhaled allergens. Furthermore, higher BMI was associated with higher tIgE, but not with sIgE. One previous study on tIgE and BMI supports our findings, though median age was lower than in our population (25). However, no former study investigated BMI as a determinant of sIgE levels in a general middle-aged and elderly population. Like smoking, higher BMI is associated with a proinflammatory state which could result in dysregulated IgE production and therefore higher tIgE levels (63, 64).

Both topical and inhaled corticosteroid use were associated with higher tIgE and sIgE levels, while oral corticosteroids showed no significant association. We found no dose-response effect of inhaled or oral corticosteroids, though this may reflect limited power due to few users. Our findings may be explained by the potential increased IgE production due to enhanced IL-4 in corticosteroid users (65-67). Interestingly, previous studies showed mixed results, with some reporting similar results and others finding lower IgE levels in corticosteroid users (65, 68-70). One explanation for this discrepancy is that we did not consider the indication of corticosteroid use, which may obscure IgE-effects. Additionally, a previous editorial suggested that IgE may temporarily rise after corticosteroid use but decline with long-term use (68). As we could not consider timing of use in our data, this may have also contributed to discrepant results.

Lastly, we found higher tIgE levels in autumn and winter compared to spring, but sIgE levels did not differ across seasons. A previous study showed no seasonal differences in tIgE (71) but suggested monthly variances in sIgE levels for some individual allergens (71, 72). Different allergens are increased across months, years and regions, which we did not consider in our analysis (73-75). Further research on season as a determinant should consider these factors and use repeated measurements to investigate intraindividual changes over time.

To our knowledge, this is the first study to investigate the determinants of tIgE and sIgE in a large population-based cohort of general middle-aged and elderly adults. Previous studies mainly focused on one or few determinants, either tIgE or sIgE, or included nongeneral populations. A strength of our study is large sample size, especially for older ages, enabling adequate statistical power to investigate the determinants of tIgE and sIgE independently. Repeated measurements allowed us to capture changes in tIgE and sIgE over time and after corticosteroid use. Unlike earlier studies, we modeled age as a continuous variable and assessed nonlinearity using splines. Moreover, we used advanced immunoassays with minimal interference for precise sIgE measurement and performed sensitivity analyses to rule out major batch or cohort effects. Our study also had potential limitations. As the Rotterdam Study primarily includes white individuals, generalizability to non-white populations may be limited. Excluding tIgE values below the detection limit in linear regression could have introduced selection bias, though Tobit regression yielded similar results, suggesting no major selection bias issue. In the longitudinal analysis, retaining undetectable tIgE values helped preserve power but may have violated model assumptions. However, diagnostics showed approximately normal residuals, suggesting minimal impact on model validity. In the longitudinal analysis, selection bias may have arisen from age differences between longitudinal and cross-sectional samples, though IPW suggested no major impact of selection bias. Additionally, longitudinal analysis of corticosteroids may have been limited by sample size and lack of consideration of timing and indication of corticosteroid use. Hence, to further clarify the longitudinal relationship between corticosteroids and IgE, future studies should research a larger sample size with more repeated measurements and incorporate indication and timing of use. Lastly, we measured sIgE for an inhaled allergen mixture, so we could not investigate determinants by allergen, which could be particularly relevant for sex, alcohol consumption and season. To draw more definitive conclusions, future studies should examine the determinants of sIgE for individual allergens.

## 6 Conclusion

We identified age, sex, smoking, BMI, season and topical and inhaled corticosteroid use as determinants of tIgE and sIgE, adding new insights into the regulation of IgE in older adults. Given the role of tIgE and sIgE as clinical biomarkers, considering these determinants in clinical practice may improve the diagnosis and management of allergic and IgE-related conditions. Future research should examine how to best incorporate these determinants into the clinical interpretation of tIgE and sIgE. Additionally, the identified determinants should be accounted for when investigating the relationship of tIgE and sIgE with chronic diseases.

## Supporting information

Supplementary tables, and legends for supplementary figures

## Data Availability

All data produced in the present study are available upon reasonable request to the authors due to privacy/ethical restrictions

## Acknowledgements

The authors are grateful to the study participants, the staff of the Rotterdam Study, and the participating general practitioners and pharmacists. We thank Hycor and Sysmex for providing support for this research.

## Tables

**Table S1:**
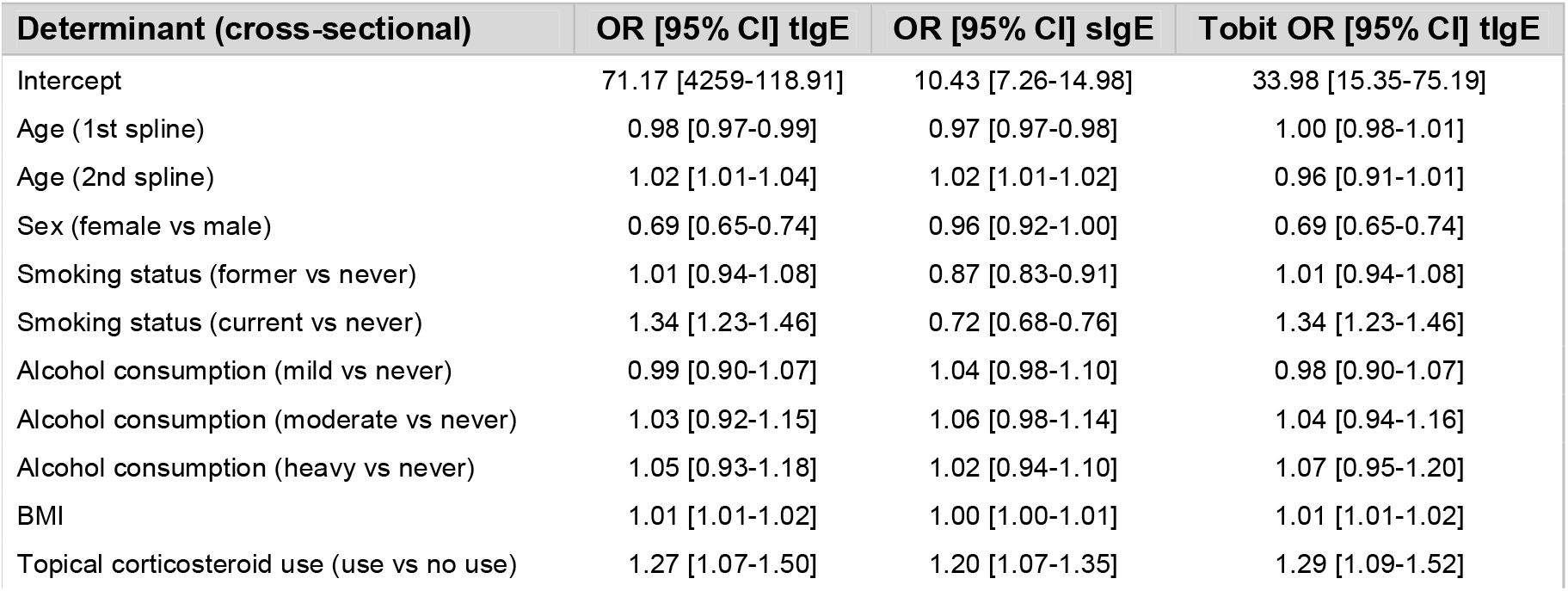
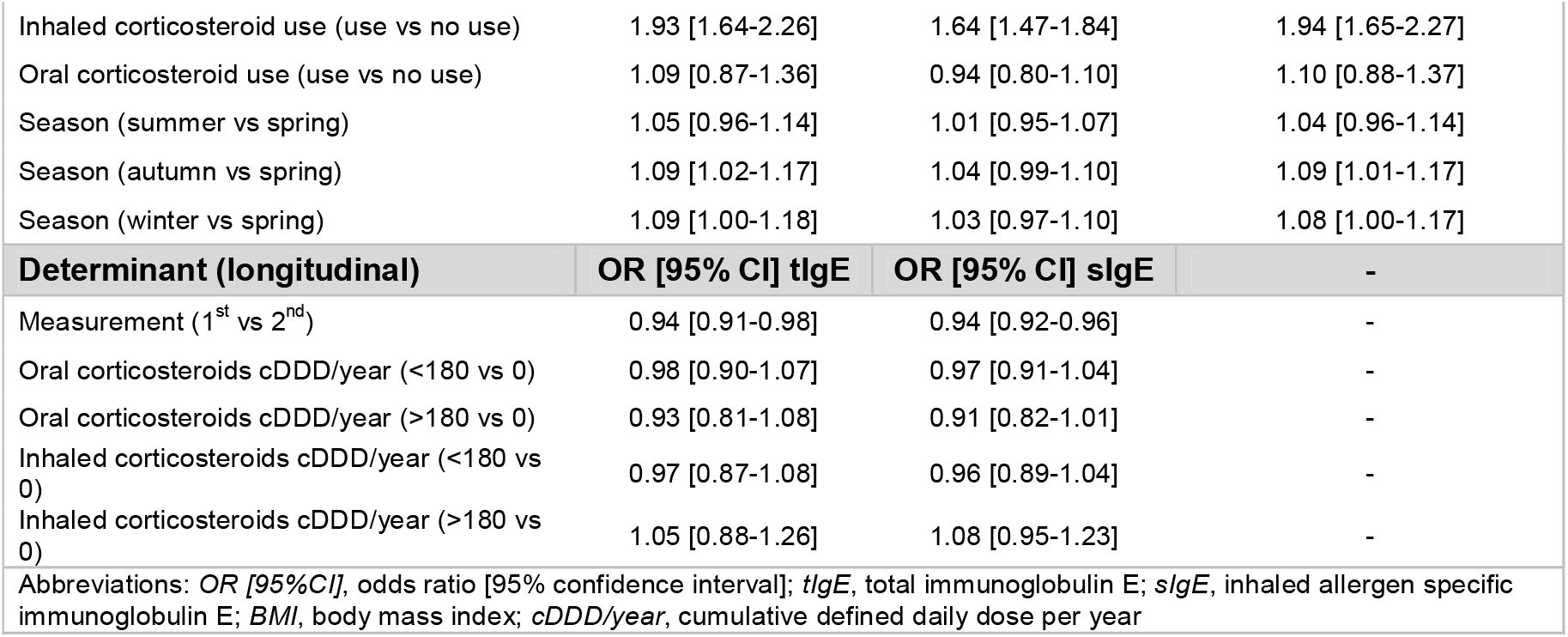
Multivariable linear regression results of tIgE and sIgE and tobit regression results of tIgE.

## References

1. Oettgen HC. Fifty years later: Emerging functions of IgE antibodies in host defense, immune regulation, and allergic diseases. The Journal of allergy and clinical immunology. 2016;137(6):1631–45.

2. Heyworth-Smith D Cp, nbsp. Laboratory Diagnosis of Allergy. 2017.

3. Siles RI, Hsieh FH. Allergy blood testing: A practical guide for clinicians. Cleveland Clinic journal of medicine. 2011;78(9):585–92.

4. Burks AW, Holgate ST, O’Hehir RE, Broide DH, Bacharier LB, Khurana Hershey GK, Peebles RS. Middleton’s allergy : principles and practice. Ninth edition. ed. Edinburgh ;: Elsevier; 2020.

5. Dorothy MJU. Negative Association between Allergy and Cancer. Scott Med J. 1969;14(2):51–4.

6. Augustin R, Chandradasa KD. IgE levels and allergic skin reactions in cancer and non-cancer patients. International archives of allergy and applied immunology. 1971;41(1):141–3.

7. Agress A, Oprea Y, Roy S, Strauch C, Rosenstreich D, Ferastraoaru D. The Association Between Malignancy, Immunodeficiency, and Atopy in IgE-Deficient Patients. The journal of allergy and clinical immunology In practice. 2024;12(1):185–94.

8. Pellizzari G, Hoskin C, Crescioli S, Mele S, Gotovina J, Chiaruttini G, et al. IgE re-programs alternatively-activated human macrophages towards pro-inflammatory anti-tumoural states. EBioMedicine. 2019;43(Journal Article):67–81.

9. Nigro EA, Brini AT, Yenagi VA, Ferreira LM, Achatz-Straussberger G, Ambrosi A, et al. Cutting Edge: IgE Plays an Active Role in Tumor Immunosurveillance in Mice. Journal of immunology (Baltimore, Md : 1950). 2016;197(7):2583–8.

10. Daniels-Wells TR, Helguera G, Leuchter RK, Quintero R, Kozman M, Rodríguez JA, et al. A novel IgE antibody targeting the prostate-specific antigen as a potential prostate cancer therapy. BMC cancer. 2013;13(Journal Article):195-.

11. Fu SL, Pierre J, Smith-Norowitz TA, Hagler M, Bowne W, Pincus MR, et al. Immunoglobulin E antibodies from pancreatic cancer patients mediate antibody-dependent cell-mediated cytotoxicity against pancreatic cancer cells. Clinical and experimental immunology. 2008;153(3):401–9.

12. Karagiannis SN, Bracher MG, Hunt J, McCloskey N, Beavil RL, Beavil AJ, et al. IgE-antibody-dependent immunotherapy of solid tumors: cytotoxic and phagocytic mechanisms of eradication of ovarian cancer cells. Journal of immunology (Baltimore, Md : 1950). 2007;179(5):2832–43.

13. Karagiannis SN, Bracher MG, Beavil RL, Beavil AJ, Hunt J, McCloskey N, et al. Role of IgE receptors in IgE antibody-dependent cytotoxicity and phagocytosis of ovarian tumor cells by human monocytic cells. Cancer immunology, immunotherapy : CII. 2008;57(2):247–63.

14. McCormick DP, Ammann AJ, Ishizaka K, Miller DG, Hong R. A study of allergy in patients with malignant lymphoma and chronic lymphocytic leukemia. Cancer. 1971;27(1):93–9.

15. Kershaw MH, Darcy PK, Trapani JA, MacGregor D, Smyth MJ. Tumor-specific IgE-mediated inhibition of human colorectal carcinoma xenograft growth. Oncology research. 1998;10(3):133–42.

16. Nagy E, Berczi I, Sehon AH. Growth inhibition of murine mammary carcinoma by monoclonal IgE antibodies specific for the mammary tumor virus. Cancer immunology, immunotherapy : CII. 1991;34(1):63–9.

17. Allegra J, Lipton A, Harvey H, Luderer J, Brenner D, Mortel R, et al. Decreased prevalence of immediate hypersensitivity (atopy) in a cancer population. Cancer research. 1976;36(9 pt.1):3225–6.

18. Xu Z, Wang T, Guo X, Li Y, Hu Y, Ma C, Wang J. The Relationship of Serum Antigen-Specific and Total Immunoglobulin E with Adult Cardiovascular Diseases. International journal of medical sciences. 2018;15(11):1098–104.

19. Sarlus H, Höglund CO, Karshikoff B, Wang X, Lekander M, Schultzberg M, Oprica M. Allergy influences the inflammatory status of the brain and enhances tau-phosphorylation. Journal of Cellular and Molecular Medicine. 2012;16(10):2401–12.

20. Pate MB, Smith JK, Chi DS, Krishnaswamy G. Regulation and dysregulation of immunoglobulin E: a molecular and clinical perspective. Clinical and molecular allergy : CMA. 2010;8(Journal Article):3-.

21. Grundbacher FJ. Causes of variation in serum IgE levels in normal populations. Journal of Allergy and Clinical Immunology. 1975;56(2):104–11.

22. Zieg G, Lack G, Harbeck RJ, Gelfand EW, Leung DY. In vivo effects of glucocorticoids on IgE production. The Journal of allergy and clinical immunology. 1994;94(2 Pt 1):222–30.

23. Settipane GA, Pudupakkam RK, McGowan JH. Corticosteroid effect on immunoglobulins. The Journal of allergy and clinical immunology. 1978;62(3):162–6.

24. Posey WC, Nelson HS, Branch B, Pearlman DS. The effects of acute corticosteroid therapy for asthma on serum immunoglobulin levels. The Journal of allergy and clinical immunology. 1978;62(6):340–8.

25. Carballo I, Alonso-Sampedro M, Gonzalez-Conde E, Sanchez-Castro J, Vidal C, Gude F, Gonzalez-Quintela A. Factors Influencing Total Serum IgE in Adults: The Role of Obesity and Related Metabolic Disorders. International archives of allergy and immunology. 2021;182(3):220–8.

26. Tokura Y. Extrinsic and intrinsic types of atopic dermatitis. Journal of dermatological science. 2010;58(1):1–7.

27. Holm Jg ATCMLTS. Determinants of disease severity among patients with atopic dermatitis: association with components of the atopic march. Archives of Dermatological Research. 2019(Journal Article):311(3):173–82.

28. Ikram MA, Brusselle GGO, Murad SD, van Duijn CM, Franco OH, Goedegebure A, et al. The Rotterdam Study: 2018 update on objectives, design and main results. European journal of epidemiology. 2017;32(9):807–50.

29. World Health O. Defined Daily Dose (DDD). 2025.

30. Hycor Biomedical I. Hycor Biomedical Inc. NOVEOS-operator’s manual. 2018(Journal Article):1–268.

31. Bauersachs D, Potapova E, Renz H, Benes SH, Matricardi PM, Skevaki C. Validation of the analytical performance of the NOVEOS™ System a system which improves upon the third-generation in vitro allergy testing technology. 2020;58(11):1865–74.

32. van Buuren S, Groothuis-Oudshoorn K. mice: Multivariate Imputation by Chained Equations in R. Journal of Statistical Software. 2011;45(3):1.

33. Venables Wn Rbd. Modern Applied Statistics with S. 2002; Fourth edition(Journal Article).

34. Arne H. censReg: Censored Regression (Tobit) Models. 2024(Journal Article).

35. Bates D, Mächler M, Bolker B, Walker S. Fitting Linear Mixed-Effects Models Using lme4. Journal of Statistical Software. 2015;67(1):1.

36. Park HJ, Kim EJ, Yoon D, Lee JK, Chang WS, Lim YM, et al. Prevalence of Self-reported Allergic Diseases and IgE Levels: A 2010 KNHANES Analysis. Allergy, asthma & immunology research. 2017;9(4):329–39.

37. De Amici M, Ciprandi G. The Age Impact on Serum Total and Allergen-Specific IgE. Allergy, asthma & immunology research. 2013;5(3):170–4.

38. Stoy PJ, Roitman-Johnson B, Walsh G, Gleich GJ, Mendell N, Yunis E, Blumenthal MN. Aging and serum immunoglobulin E levels, immediate skin tests, RAST. The Journal of allergy and clinical immunology. 1981;68(6):421–6.

39. Barbee RA, Halonen M, Lebowitz M, Burrows B. Distribution of IgE in a community population sample: correlations with age, sex, and allergen skin test reactivity. Journal of Allergy and Clinical Immunology. 1981;68(2):106–11.

40. Grigoreas C, Pappas D, Galatas ID, Kollias G, Papadimos S, Papadakis P. Serum total IgE levels in a representative sample of a Greek population. I. Correlation with age, sex, and skin reactivity to common aeroallergens. Allergy. 1993;48(3):142–6.

41. Holford-Strevens V, Warren P, Wong C, Manfreda J. Serum total immunoglobulin E levels in Canadian adults. The Journal of allergy and clinical immunology. 1984;73(4):516–22.

42. Barbee RA, Halonen M, Kaltenborn W, Lebowitz M, Burrows B. A longitudinal study of serum IgE in a community cohort: correlations with age, sex, smoking, and atopic status. The Journal of allergy and clinical immunology. 1987;79(6):919–27.

43. Kerkhof M, Droste JH, de Monchy JG, Schouten JP, Rijcken B. Distribution of total serum IgE and specific IgE to common aeroallergens by sex and age, and their relationship to each other in a random sample of the Dutch general population aged 20-70 years. Dutch ECRHS Group, European Community Respiratory Health Study. Allergy. 1996;51(11):770–6.

44. Amaral AFS, Newson RB, Abramson MJ, Antó JM, Bono R, Corsico AG, et al. Changes in IgE sensitization and total IgE levels over 20 years of follow-up. The Journal of allergy and clinical immunology. 2016;137(6):1788-95.e9.

45. Vasto S, Malavolta M, Pawelec G. Age and immunity. Immunity & ageing : I & A. 2006;3(Journal Article):2-.

46. Khan SR, van der Burgh AC, Peeters RP, van Hagen PM, Dalm Vash, Chaker L. Determinants of Serum Immunoglobulin Levels: A Systematic Review and Meta-Analysis. Frontiers in Immunology. 2021;olume 12-2021(Journal Article).

47. Khan SR, Chaker L, Ikram MA, Peeters RP, van Hagen PM, Dalm VASH. Determinants and Reference Ranges of Serum Immunoglobulins in Middle-Aged and Elderly Individuals: a Population-Based Study. Journal of clinical immunology. 2021;41(8):1902–14.

48. Leffler J, Stumbles PA, Strickland DH. Immunological Processes Driving IgE Sensitisation and Disease Development in Males and Females. International journal of molecular sciences. 2018;19(6):1554. doi: 10.3390/ijms19061554.

49. Jarvis D, Luczynska C, Chinn S, Burney P. The association of age, gender and smoking with total IgE and specific IgE. Clinical and experimental allergy : journal of the British Society for Allergy and Clinical Immunology. 1995;25(11):1083–91.

50. Omenaas E, Bakke P, Elsayed S, Hanoa R, Gulsvik A. Total and specific serum IgE levels in adults: relationship to sex, age and environmental factors. Clinical and experimental allergy : journal of the British Society for Allergy and Clinical Immunology. 1994;24(6):530–9.

51. Roved J, Westerdahl H, Hasselquist D. Sex differences in immune responses: Hormonal effects, antagonistic selection, and evolutionary consequences. Hormones and behavior. 2017;88(Journal Article):95–105.

52. Jensen-Jarolim E, Untersmayr E. Gender-medicine aspects in allergology. Allergy. 2008;63(5):610–5.

53. Warren CPW, Holford-Strevens V, Wong C, Manfreda J. The relationship between smoking and total immunoglobulin E levels. Journal of Allergy and Clinical Immunology. 1982;69(4):370–5.

54. Swetha M C M M H. Assessment of serum total IGE levels in smokers, non smokers and ex smokers and its relation to lung function, airway symptoms and atopic predisposition. IP Indian Journal of Immunology and Respiratory Medicine. 2019;2019(Journal Article):10227.

55. Kim YS, Kim HY, Ahn H-S, Sohn TS, Song JY, Lee YB, et al. The Association between Tobacco Smoke and Serum Immunoglobulin E Levels in Korean Adults. Internal medicine (Tokyo, Japan). 2017;56(19):2571–7.

56. Bahna SL, Heiner DC, Myhre BA. Immunoglobulin E pattern in cigarette smokers. Allergy. 1983;38(1):57–64.

57. le floch-ramondou a, Papatheodorou A, Scott G, Asrat S, Murphy A, Sleeman M, Orengo J. erj. p. PA4402.

58. Byron KA, Varigos GA, Wootton AM. IL-4 production is increased in cigarette smokers. Clinical and experimental immunology. 1994;95(2):333–6.

59. Wüthrich B, Schindler C, Medici TC, Zellweger JP, Leuenberger P. IgE levels, atopy markers and hay fever in relation to age, sex and smoking status in a normal adult Swiss population. SAPALDIA (Swiss Study on Air Pollution and Lung Diseases in Adults) Team. International archives of allergy and immunology. 1996;111(4):396–402.

60. Lomholt FK, Nielsen SF, Nordestgaard BG. High alcohol consumption causes high IgE levels but not high risk of allergic disease. Journal of Allergy and Clinical Immunology. 2016;138(5):1404-13.e13.

61. Roh D, Lee D-H, Lee S-K, Kim SW, Kim SW, Cho JH, et al. Sex difference in IgE sensitization associated with alcohol consumption in the general population. Scientific Reports. 2019;9(1):12131.

62. Vidal C, Armisén M, Domínguez-Santalla MJ, Gude F, Lojo S, González-Quintela A. Influence of alcohol consumption on serum immunoglobulin E levels in atopic and nonatopic adults. Alcoholism, Clinical and Experimental Research. 2002;26(1):59–64.

63. Ellulu MS, Patimah I, Khaza’ai H, Rahmat A, Abed Y. Obesity and inflammation: the linking mechanism and the complications. Archives of medical science : AMS. 2017;13(4):851–63.

64. Hersoug LG, Linneberg A. The link between the epidemics of obesity and allergic diseases: does obesity induce decreased immune tolerance? Allergy. 2007;62(10):1205–13.

65. Salvi SS, Babu KS, Holgate ST. Glucocorticoids enhance IgE synthesis. Are we heading towards new paradigms? Clinical & Experimental Allergy. 2000;30(11):1499–505.

66. Jabara HH, Ahern DJ, Vercelli D, Geha RS. Hydrocortisone and IL-4 induce IgE isotype switching in human B cells. Journal of immunology (Baltimore, Md : 1950). 1991;147(5):1557–60.

67. Jabara HH, Brodeur SR, Geha RS. Glucocorticoids upregulate CD40 ligand expression and induce CD40L-dependent immunoglobulin isotype switching. The Journal of clinical investigation. 2001;107(3):371–8.

68. Corne J. Do inhaled corticosteroids reduce serum IgE levels? The answer is maybe but how relevant is the question? Clinical and experimental allergy : journal of the British Society for Allergy and Clinical Immunology. 1999;29(3):294–7.

69. Ohrui T, Funayama T, Sekizawa K, Yamaya M, Sasaki H. Effects of inhaled beclomethasone dipropionate on serum IgE levels and clinical symptoms in atopic asthma. Clinical and experimental allergy : journal of the British Society for Allergy and Clinical Immunology. 1999;29(3):357–61.

70. Diaz-Sanchez D, Tsien A, Fleming J, Saxon A. Effect of topical fluticasone propionate on the mucosal allergic response induced by ragweed allergen and diesel exhaust particle challenge. Clinical immunology (Orlando, Fla). 1999;90(3):313–22.

71. Lam HCY, Jarvis D, the Ecrhs II. Seasonal variation in total and pollen-specific immunoglobulin E levels in the European Community Respiratory Health Survey. Clinical & Experimental Allergy. 2021;51(8):1085–8.

72. Beeh KM, Beier J, Buhl R. Seasonal variations of serum-IgE and potential impact on dose-calculation of omalizumab (rhuMab-E25, anti-IgE). Pneumologie (Stuttgart, Germany). 2004;58(8):546–51.

73. Lo F, Bitz CM, Battisti DS, Hess JJ. Pollen calendars and maps of allergenic pollen in North America. Aerobiologia. 2019;35(4):613–33.

74. Lake IR, Jones NR, Agnew M, Goodess CM, Giorgi F, Hamaoui-Laguel L, et al. Climate Change and Future Pollen Allergy in Europe. Environmental health perspectives. 2017;125(3):385–91.

75. Tham EH, Lee AJ, Bever HV. Aeroallergen sensitization and allergic disease phenotypes in Asia. Asian Pacific journal of allergy and immunology. 2016;34(3):181–9.

