## Supplementary tables, and legends for supplementary figures for "Determinants of total and inhaled allergen-specific immunoglobulin E in the middle-aged and elderly population"

**Supplementary tables and figure legends**

| **Table S1: Multivariable linear regression results of tIgE and sIgE and tobit regression results of tIgE** | | | |
| --- | --- | --- | --- |
| **Determinant (cross-sectional)** | **OR [95% CI] tIgE** | **OR [95% CI] sIgE** | **Tobit OR [95% CI] tIgE** |
| Intercept | 71.17 [4259-118.91] | 10.43 [7.26-14.98] | 33.98 [15.35-75.19] |
| Age (1st spline) | 0.98 [0.97-0.99] | 0.97 [0.97-0.98] | 1.00 [0.98-1.01] |
| Age (2nd spline) | 1.02 [1.01-1.04] | 1.02 [1.01-1.02] | 0.96 [0.91-1.01] |
| Sex (female vs male) | 0.69 [0.65-0.74] | 0.96 [0.92-1.00] | 0.69 [0.65-0.74] |
| Smoking status (former vs never) | 1.01 [0.94-1.08] | 0.87 [0.83-0.91] | 1.01 [0.94-1.08] |
| Smoking status (current vs never) | 1.34 [1.23-1.46] | 0.72 [0.68-0.76] | 1.34 [1.23-1.46] |
| Alcohol consumption (mild vs never) | 0.99 [0.90-1.07] | 1.04 [0.98-1.10] | 0.98 [0.90-1.07] |
| Alcohol consumption (moderate vs never) | 1.03 [0.92-1.15] | 1.06 [0.98-1.14] | 1.04 [0.94-1.16] |
| Alcohol consumption (heavy vs never) | 1.05 [0.93-1.18] | 1.02 [0.94-1.10] | 1.07 [0.95-1.20] |
| BMI | 1.01 [1.01-1.02] | 1.00 [1.00-1.01] | 1.01 [1.01-1.02] |
| Topical corticosteroid use (use vs no use) | 1.27 [1.07-1.50] | 1.20 [1.07-1.35] | 1.29 [1.09-1.52] |
| Inhaled corticosteroid use (use vs no use) | 1.93 [1.64-2.26] | 1.64 [1.47-1.84] | 1.94 [1.65-2.27] |
| Oral corticosteroid use (use vs no use) | 1.09 [0.87-1.36] | 0.94 [0.80-1.10] | 1.10 [0.88-1.37] |
| Season (summer vs spring) | 1.05 [0.96-1.14] | 1.01 [0.95-1.07] | 1.04 [0.96-1.14] |
| Season (autumn vs spring) | 1.09 [1.02-1.17] | 1.04 [0.99-1.10] | 1.09 [1.01-1.17] |
| Season (winter vs spring) | 1.09 [1.00-1.18] | 1.03 [0.97-1.10] | 1.08 [1.00-1.17] |
| **Determinant (longitudinal)** | **OR [95% CI] tIgE** | **OR [95% CI] sIgE** | **-** |
| Measurement (1^st^ vs 2^nd^) | 0.94 [0.91-0.98] | 0.94 [0.92-0.96] | - |
| Oral corticosteroids cDDD/year (<180 vs 0) | 0.98 [0.90-1.07] | 0.97 [0.91-1.04] | - |
| Oral corticosteroids cDDD/year (>180 vs 0) | 0.93 [0.81-1.08] | 0.91 [0.82-1.01] | - |
| Inhaled corticosteroids cDDD/year (<180 vs 0) | 0.97 [0.87-1.08] | 0.96 [0.89-1.04] | - |
| Inhaled corticosteroids cDDD/year (>180 vs 0) | 1.05 [0.88-1.26] | 1.08 [0.95-1.23] | - |
| Abbreviations: *OR [95%CI]*, odds ratio [95% confidence interval]; *tIgE*, total immunoglobulin E; *sIgE*, inhaled allergen specific immunoglobulin E; *BMI*, body mass index; *cDDD/year*, cumulative defined daily dose per year | | | |

| **Table S2: Population characteristics in all cohorts, ERGOjong and the longitudinal sample** | | | |
| --- | --- | --- | --- |
|  | **Cross-sectional:**  **All cohorts (N=8679)** | **Cross-sectional: ERGOjong (n= 3443)** | **Longitudinal (n=478)** |
| **Variable** | ***n* (%), unless stated otherwise** | ***n* (%), unless stated otherwise** | ***n* (%), unless stated**  **otherwise** |
| Age, *years,* mean (SD) | 64.18 (9.82) | 57.10 (6.83) | 57.02 (5.77) |
| *Years,* Range | 45.57 - 105.78 | 45.57 - 97.32 | 46.56 - 84.19 |
| Sex (female) | 4941 (56.9) | 1943 (56.4) | 276 (57.7) |
| Smoking status |  |  |  |
| Never | 2880 (33.2) | 1106 (32.1) | 160 (33.5) |
| Former | 4106 (47.3) | 1542 (44.8) | 229 (47.9) |
| Current | 1693 (19.5) | 795 (23.1) | 89 (18.6) |
| Alcohol consumption^a^ |  |  |  |
| Never | 1624 (18.7) | 371 (10.8) | 43 (9.0) |
| Mild (<10 g/day) | 4528 (52.2) | 2214 (64.3) | 322 (67.4) |
| Moderate (10-20 g/day) | 1525 (17.6) | 632 (18.4) | 88 (18.4) |
| Heavy (>20 g/day) | 1002 (11.5) | 226 (6.6) | 25 (5.2) |
| BMI, *kg/m²,* mean (SD) | 27.33 (4.26) | 27.1 (4.61) | 27.34 (4.45) |
| *Kg/m²*, Range | 12.62 - 56.87 | 12.62 - 56.87 | 17.15 - 50.21) |
| Oral corticosteroid use (yes) | 154 (1.8) | 30 (0.9) | 112 (23.4)^b^ |
| 0 cDDD/year | - | - | 366 (76.6) |
| <180 cDDD/year | - | - | 85 (17.8) |
| >180 cDDD/year | - | - | 27 (5.6) |
| Inhaled corticosteroid use (yes) | 281 (3.2) | 119 (3.5) | 70 (14.7)^b^ |
| 0 cDDD/year | - | - | 408 (85.4) |
| <180 cDDD/year | - | - | 52 (10.9) |
| >180 cDDD/year | - | - | 18 (3.8) |
| Topical corticosteroid use (yes) | 276 (3.2) | 70 (2.0) | 10 (2.1) |
| tIgE, *kU/L*, median [IQR] | 24.30 [9.97 - 71.2] | 25.1 [9.92 - 75.5] | 22.95 [9.29 - 76.5] |
| tIgE, *kU/L*, Range | 5-5000^c^ | 5-5000 | 5-5000^c^ |
| tIgE (<5) | 920 (10.6) | 368 (10.7) | 52 (10.9) |
| tIgE (5-100) | 6075 (70.0) | 2368 (68.8) | 324 (67.8) |
| tIgE (>100) | 1676 (19.3) | 702 (20.4) | 102 (21.3) |
| tIgE (value missing) | 8 (0.1) | 5 (0.1) | - |
| sIgE, *SAU*, median [IQR] | 0.12 [0.08 - 0.20] | 0.12 [0.08 - 0.53] | 0.10 [0.07 - 0.67] |
| sIgE, *SAU*, Range | 0.00 - 426.22 | 0 – 426.22 | 0.03 - 349.44 |
| sIgE (<1) | 7228 (83.3) | 2656 (77.1) | 363 (75.9) |
| sIgE (>1) | 1434 (16.5) | 781 (22.7) | 115 (24.1) |
| sIgE (value missing) | 17 (0.2) | 6 (0.2) | - |
| Season | 2786 (32.1) | 1123 (32.6) | 138 (28.9) |
| Spring | 2786 (32.1) | 1123 (32.6) | 138 (28.9) |
| Summer | 1506 (17.4) | 1063 (30.9) | 83 (17.4) |
| Autumn | 2642 (30.4) | 624 (18.1) | 156 (32.6) |
| Winter | 1745 (20.1) | 633 (18.4) | 101 (21.1) |
| Follow-up time, *years,* median [IQR] | - | - | 5.56 [5.42 - 5.78] |
| ^a^Categories according to the World Health Organization guidelines of alcohol consumption  ^b^Total participants that used corticosteroids between the 1st and 2nd measurement  ^c^5 and 5000 kU/L are the detection limits. Values below 5 are measured as 5 and values above 5000 are measured as 5000 | | | |
| Abbreviations: *SD*, standard deviation; *BMI*, body mass index; *cDDD/year*, cumulative defined daily dose per year; *tIgE*, total immunoglobulin E; *kU/L*, kilounits per liter; *IQR*, interquartile range; *sIgE*, inhaled allergen specific immunoglobulin E; *SAU*, Standard Arbitrary Units | | | |

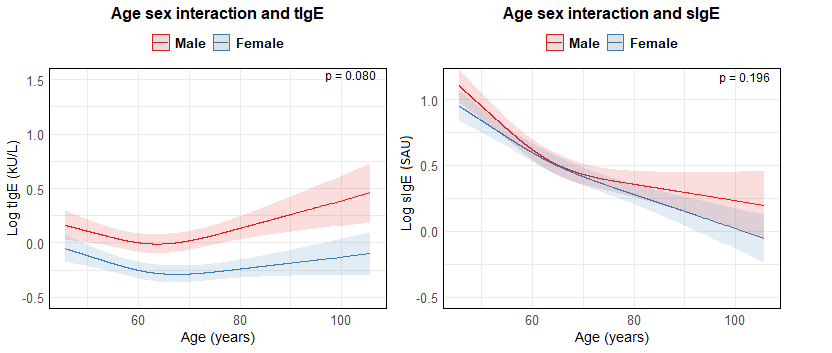
**Figure S1:** Age-sex interaction for tIgE and sIgE

**Abbreviations**: *tIgE*, total immunoglobulin E, *kU/L*, kilounits per liter; s*IgE*, inhaled allergen specific immunoglobulin E; *SAU*, standard arbitrary units

**Description**: **Figure S1** shows the age-sex interaction for tIgE on the left and for sIgE on the right. Both models were adjusted for age, sex, smoking, alcohol consumption, BMI, season and topical, inhaled and oral corticosteroids. The tIgE values were log-transformed after exclusion of values below the detection limit. The sIgE values were log-transformed after exclusion of zero values. The effect estimates are standardized through z-score standardization and plotted as beta coefficient per standard deviation.

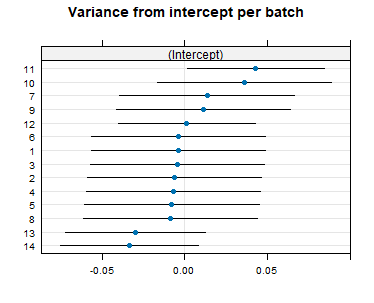

**Figure S2:** Variance from the intercept per batch for the sIgE model

**Description**: The linear mixed model with batch as clustering variable did not suggest relevant batch differences (intraclass correlation, ICC: 0.002). As depicted in figure 3, sIgE values were higher in batch 10 and 11 compared to the reference batch 1, but excluding those batches did not change the results meaningfully.
